# High-Risk Anti-Seizure Medication Use in Childbearing-Age People with Epilepsy in a *Taenia solium* Endemic Region

**DOI:** 10.64898/2026.06.08.26354652

**Authors:** Samantha E. Allen, Melissa T. Wardle, Luz M. Moyano, Percy Vilchez, Javier A. Bustos, Hector H. Garcia, Seth E. O’Neal, Cysticercosis Working Group in Peru (CWGP)

**Affiliations:** University of California, Davis, Department of Neurology, 3160 Folsom Blvd., Suite 2100. Sacramento, CA 95817 USA; Oregon Health and Science University – Portland State University, School of Public Health, 810 SW 5th Ave, Portland, OR 97201 USA; Universidad Nacional de Tumbes, School of Medicine, Av. Universitaria S/N - Pampa Grande Tumbes, Tumbes 24001 Peru; Universidad Peruana Cayetano Heredia, Centro de Salud Global, Tumbes, Peru; Instituto Nacional de Ciencias Neurologicas, Jr. Ancash 1271 Barrios Altos, Lima 15003, Peru; Universidad Peruana Cayetano Heredia, Department of Microbiology, School of Sciences, Avenida Honorio Delgado 430, Urb. Ingeniería, San Martín de Porres, Lima 15102, Peru

## Abstract

**Background/Objectives:** People of childbearing potential with epilepsy who live in regions endemic for *Taenia solium* represent a particularly vulnerable population due to the high burden of epilepsy related to neurocysticercosis (NCC) combined with limited access to safer anti-seizure medications (ASMs). However, the extent of these resource limitations and current prescribing practices remains poorly characterized, limiting the development of appropriate clinical guidelines. This study aimed to characterize ASM prescribing patterns, including pregnancy-associated risk profiles and polytherapy, and to evaluate how these patterns vary by demographic and clinical characteristics among people of childbearing potential in an epilepsy cohort in Peru, a region highly endemic for *T. solium*.

**Methods:** Participants were drawn from a prospective, population-based epilepsy cohort established in Tumbes, Peru (2006–2020). The analytic population included individuals with epilepsy of childbearing potential, defined as females aged 15–49 years. The primary outcome was ASM risk, categorized by pregnancy-associated risk of congenital malformations and adverse neurodevelopmental outcomes. ASMs were classified as “Established Low Risk” (lamotrigine, levetiracetam), “Possible Risk/Inadequate Data” (carbamazepine, phenobarbital, phenytoin), and “Established High Risk” (valproic acid). Prescription patterns were analyzed in relation to demographic and clinical characteristics.

**Results:** Among 1,975 individuals with epilepsy in the cohort, 685 were people of childbearing potential. Approximately one-third met criteria for probable or definite NCC (34.9%). Most ASM prescriptions fell into the “Possible Risk/Inadequate Data” category (87.0%), while 12.8% of participants received medications in the “Established High Risk” category. In multivariable analysis, factors associated with high-risk prescriptions included ASM use prior to enrollment and polytherapy.

**Discussion:** People of childbearing potential with epilepsy in this cohort were predominantly treated with carbamazepine, phenytoin, phenobarbital, and valproate, the ASMs most consistently available in this region during the study period. Despite strong evidence supporting the use of newer-generation agents such as lamotrigine and levetiracetam for individuals who may become pregnant, prescribing patterns in this cohort reflect significant formulary and resource constraints. Current recommendations for the management of people of childbearing potential with epilepsy do not adequately address the realities of care in resource-limited, NCC-endemic settings.

## Introduction

Epilepsy affects an estimated 50 million people worldwide, with a disproportionate burden in low- and middle-income countries (LMICs).^1^ In regions endemic for *Taenia solium*, neurocysticercosis (NCC) or *T. solium* infection of the central nervous system, represents one of the leading causes of acquired epilepsy.^2^ For those afflicted, effective long-term seizure control remains a critical challenge, particularly in resource-limited settings where these infections are prevalent and access to anti-seizure medications (ASMs) is often constrained.^3^

Among those impacted, people of childbearing potential with epilepsy represent a particularly vulnerable population that deserves special consideration. ASM choice in this group must balance seizure control with teratogenic risk and other potential adverse maternal and fetal outcomes. In determining ASM use in this population, guidelines recommend individualized therapy with preference for safer ASMs like levetiracetam and lamotrigine,^4–6^ together with preconception counseling; however, these safer ASMs are typically unavailable, and access to preconception services are limited in LMICs.^5,7^ Undoubtedly, LMICs need guidelines that consider resource limitations. Unfortunately, specificity of guidelines is limited by a paucity of data on ASM use and prescription patterns for people of childbearing potential in LMICs, particularly in populations with the further burden of NCC, where the burden of epilepsy may be higher and treatment gaps more pronounced.^8^

Indeed, across LMICs, there is a dramatic epilepsy treatment gap in terms of access, diagnostic and therapeutic capabilities, quality of care and other unmet health needs. Systematic reviews have estimated that more than 75% of individuals with epilepsy do not receive appropriate treatment, and access is especially poor in rural settings.^8,9^ This is often due to limited availability of ASMs, high cost, inconsistent supply, and structural barriers in health systems.^10^

In low resource settings, prescription patterns are constrained by available ASMs and thus favor older, lower-cost drugs, such as phenobarbital, phenytoin, carbamazepine, and valproate, rather than newer agents with potentially better safety or tolerability profiles and lower risk of teratogenicity.^10^ In a recent systematic review in LMICs, the majority of studies (82.3%) reported use of first-generation ASMs, with limited adoption of newer medications.^11,12^

Northern Peru is highly endemic for *T. solium*, and a large, population-based cohort was established to study epilepsy in this region.^13^ This cohort has been previously leveraged to describe baseline ASM use, quality of life, and the epidemiology of NCC-associated epilepsy. However, ASM use among people of childbearing potential in this setting has not been systematically examined, representing a critical gap in understanding the intersection of epilepsy management, reproductive health, and NCC burden.

This study aims to characterize the patterns of ASM prescribing, including specific ASM risk profiles and polytherapy, and how these patterns vary by participant characteristics, among people of childbearing potential in the Northern Peru epilepsy cohort. Understanding ASM use in this high-risk population has important implications for clinical management, reproductive counseling, and public health interventions in NCC-endemic regions.

## Methods

### Study Design and Setting

Participants were drawn from a prospective, population-based epilepsy cohort established in Tumbes, Northern Peru, a region highly endemic for *T. solium*. The cohort was designed to evaluate epilepsy epidemiology, clinical characteristics, and outcomes in both people with and without NCC.^13^ Enrollment occurred between 2006 and 2020 through community-based screening in 107 villages, home visits, and self-referrals to local clinics. Participants underwent standardized clinical evaluations, including neurologic assessment, neuroimaging, serologic testing, and electroencephalography (EEG).

### Study Population

Inclusion criteria for this analysis were: (1) diagnosis of epilepsy, defined as two or more unprovoked seizures separated by at least 24 hours; (2) residence in a *T. solium*–endemic region; and (3) being of childbearing potential, defined as individuals identified as biologically female and aged 15-49 years, consistent with the World Health Organization (WHO) defined reproductive age range.^14^ Participants without epilepsy or outside the reproductive age range were excluded. Participants identified as male within the same reproductive age range (15–49 years) were included in a secondary analysis as a comparator group to contextualize ASM prescribing patterns in the overall population.

### Data Collection

Demographic, clinical, and treatment information was collected using structured questionnaires administered by trained research personnel. ASM use was recorded, including type, dose, and use of concurrent medications at the time of baseline intake. ASM treatment decisions were tailored to individual patient circumstances and left to the discretion of the treating physician. ASM selection was in no way impacted by the cohort study procedures. Additional data collected included year of recruitment, history of developmental delay, marital status, contraception use, seizure type, duration of epilepsy, and NCC diagnostic status using the modified Del Brutto criteria.^15^

### ASM Classification

ASMs were categorized as first-generation (e.g., phenobarbital, phenytoin, carbamazepine, and valproate) or newer generation (e.g., levetiracetam, lamotrigine). ASMs were also categorized by their pregnancy-associated risk of fetal malformations and adverse neurodevelopmental outcomes to define the primary outcome, ASM risk-profile. ASMs were classified as “Established Low Risk” (lamotrigine and levetiracetam), “Possible Risk/Inadequate Data” (carbamazepine, oxcarbazepine, phenobarbital, phenytoin, topiramate, and zonisamide), and “Established High Risk” (valproic acid).^16–20^ Given sparse observations in the low-risk category, these medications were classified with the intermediate Possible Risk/Inadequate Data group for analysis. Polytherapy was defined as the use of two or more ASMs concurrently. ASMs considered reliably available in this region of Peru during the study period (2006 through 2020) include phenobarbital, phenytoin, carbamazepine, and valproic acid, which are included on the national essential medicines list.

### Statistical Analysis

Descriptive statistics are used to summarize demographic and clinical characteristics. Categorical variables are expressed as counts and percentages; continuous variables as means with standard deviation (SD). Prescription patterns were analyzed in relation to reproductive potential, NCC status, seizure type, and participant demographic characteristics. Group comparisons were performed with Chi-square or Fisher’s exact tests for categorical variables; independent t-tests for continuous variables. Multivariable logistic regression was used to evaluate factors associated with the use of “established high risk” ASMs among those prescribed ASMs. ASM risk-profile was used as the primary dependent variable and adjusting for age, marital status, epilepsy type among those with a known epilepsy classification (generalized or focal), use of ASM prior to enrollment, and number of ASMs prescribed. These variables were selected based on the results of the univariate analysis (p < 0.10) and known associates established in the literature.^21,22^ Descriptive analyses were conducted using all available observations for each variable, while regression statistical analyses, including multivariable regression models, were restricted to complete cases for included covariates. A significance threshold of p < 0.05 was used. Statistical analyses were performed using Stata version 19 (StataCorp, College Station, TX).

## Results

### Cohort Characteristics

Out of the full cohort of individuals with epilepsy totaling 1975 participants, there were a total of 685 biologically female (people of childbearing potential) from age 15 to 49. Mean age was 30.1 years (SD 9.9). A little over half reported “married” as their marital status (52.6%) and of those who reported on contraception use (n=223), 32% reported no use of contraception, 19% long-acting highly effective contraception (IUD or surgical), 44% hormonal (pill or injectable), and a minority 5% using condoms. Approximately 10.5% had some history of developmental delay. Focal seizures predominated (60%) with a mean duration of diagnosis of 13.8 years (SD 11.5). A total of 239 participants (34.9%) met criteria for probable or definite NCC (Table 1), and the majority of cases had calcified lesions only (84%) (data not shown).

**Table 1:** Baseline characteristics of cohort participants of childbearing potential.

|  | No.* (%) |
| --- | --- |
| **Participants aged 15-49** | 685 |
| **Average age (SD)** | 30.1 (9.9) |
| **Married marital status** | 360 (52.6) |
| **History of developmental delay** | 72 (10.5) |
| **Method of contraception** |  |
| None | 74 (10.8) |
| Tubal ligation/surgical | 34 (5.0) |
| Oral Hormonal | 33 (4.8) |
| Injectable Hormonal | 71 (10.4) |
| Intrauterine Device | 11 (1.6) |
| Condoms | 11 (1.6) |
| Missing | 451 (65.8) |
| **Epilepsy Type** |  |
| Generalized | 116 (16.9) |
| Focal | 411 (60.0) |
| Unknown | 12 (1.8) |
| Missing | 146 (21.3) |
| **NCC Diagnosis** | 239 (34.9) |
| **Duration of epilepsy** |  |
| 0-10 years | 272 (39.7) |
| >10 years | 310 (45.3) |
| Missing | 103 (15.0) |
| **ASM prior to recruitment** | 162 (23.6) |
| **Number of ASMs prescribed at baseline** |  |
| None | 138 (20.2) |
| Monotherapy | 487 (71.1) |
| Polytherapy | 60 (8.8) |
| **First Generation ASM prescription (N=547)** | 538 (98.6) |
| **ASM risk profile (N=547)** |  |
| Low Risk | 1 (0.2) |
| Possible Risk/Inadequate Data | 476 (87.0) |
| High Risk | 70 (12.8) |

\*Proportions are presented out of 685 unless otherwise specified. Missingness was observed for the following variables: marital status (3.6%), history of developmental delay (21.9%), and NCC diagnosis (9.1%).

### ASM Use

Among people of childbearing potential, approximately 80% (547/685) were prescribed at least one ASM at the time of baseline recruitment. Among those prescribed an ASM, monotherapy was used in 487 (89.0%) and polytherapy in 60 (11.0%). Almost all participants were prescribed first-generation ASMs (98.6%) with rare instances of newer-generation ASMs prescribed. Most baseline prescriptions fell into the “Possible Risk/Inadequate Data” category (87%) with 12.8% receiving “Established High Risk” category prescriptions (prescription counts by individual ASM in Suppl Table 2). ASM prescription patterns were evaluated based on year of enrollment from 2006 to 2020. On average, there were approximately 40 participants (range 2-81) in this population enrolled per year with peak enrollment occurring between 2012 and 2014. The percentage of participants receiving high-risk ASM prescriptions was relatively stable over the course of the enrollment period sampled with a median of 12.3% of participants in this population receiving high-risk prescriptions each year (Figure 1).

**Figure 1:**
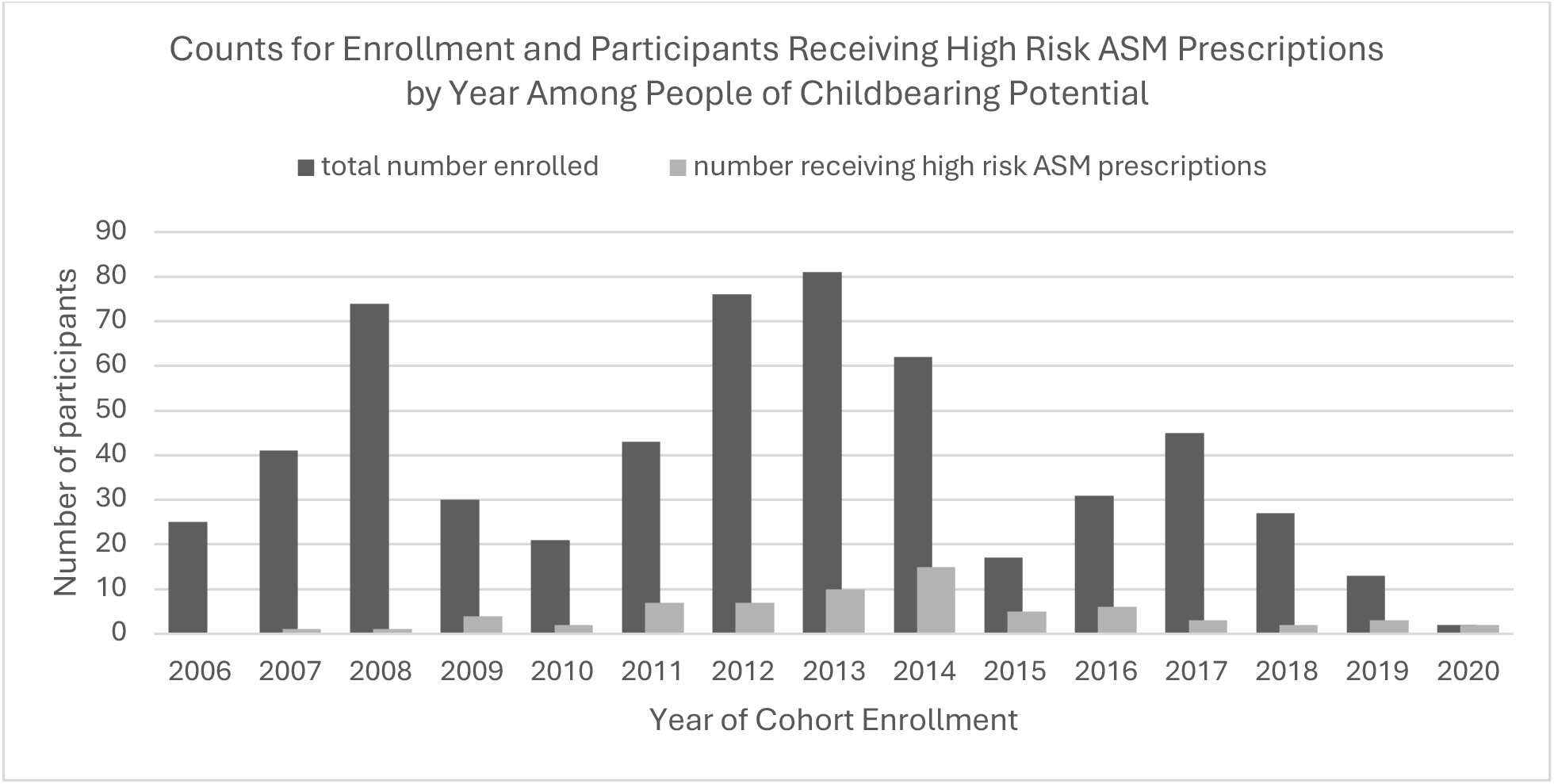
Number of High Risk ASM Prescriptions by Year of Enrollment

### Factors Associated with Use of High-Risk ASM prescriptions

In the univariate model, marital status, ASM prescriptions prior to cohort enrollment, and number of ASMs prescribed at the time of enrollment were all significantly associated with ASM risk profile (Table 2). In the multivariable logistic regression model, established high risk ASM prescriptions were associated with history of ASM prescription prior to cohort enrollment (OR 3.10, 95% CI 1.30, 7.34) and polytherapy (OR 3.89, 95% CI 1.62, 9.35) among participants prescribed ASMs after adjusting for age, marital status, and epilepsy type. No relationship was seen between marital status, age, and epilepsy type with ASM risk profile in the adjusted model (Table 3). Due to missingness, the logistic regression was limited to a population of 211 participants. These 211 participants were more likely to have focal epilepsy, a shorter duration of epilepsy, have received an ASM prescription prior to recruitment, polytherapy prescription at baseline intake, later generation ASMs prescriptions, and also more likely to be prescribed a high-risk category ASM when compared to those excluded (Table S3).

**Table 2.** Factors Associated with High-Risk ASM Use.

|  | Risk – n (%) |  | P value |
| --- | --- | --- | --- |
|  | Medium | High |  |
| **Participants aged 15-49** | 549 | 70 |  |
| **Average age (SD)** | 29.9 (9.9) | 27.7 (9.6) | 0.09 |
| **Married marital status** | 288 (54.8) | 27 (38.6) | **0.01** |
| **History of developmental delay** | 57 (13.4) | 9 (16.1) | 0.59 |
| **Use of Contraception** | 129 (67.9) | 10 (52.6) | 0.18 |
| **Epilepsy Type** |  |  |  |
| Generalized | 79 (18.6) | 15 (23.1) | 0.60 |
| Focal | 337 (79.3) | 48 (73.9) |  |
| Unknown | 9 (2.1) | 2 (3.1) |  |
| **NCC Diagnosis** | 190 (37.9) | 23 (38.3) | 0.94 |
| **Duration of epilepsy** |  |  |  |
| 0-10 years | 167 (36.1) | 19 (30.7) | 0.99 |
| >10 years | 446 (49.3) | 57 (50.4) |  |
| **ASM prior to recruitment** | 127 (50.2) | 35 (77.8) | **0.001** |
| **No. of ASMs prescribed at baseline** |  |  |  |
| None | 72 (13.1) | 0 | **<0.001** |
| Monotherapy | 440 (80.2) | 47 (67.1) |  |
| Polytherapy | 37 (6.7) | 23 (32.9) |  |

**Table 3:** Associates of High Risk ASM Use by Multivariable Logistic Regression (n=211)

|  | Odds Ratio | Standard Error | p-value | Lower 95% CI | Upper 95% CI |
| --- | --- | --- | --- | --- | --- |
| **Age** |  |  |  |  |  |
|  | 0.969 | 0.021 | 0.148 | 0.928 | 1.011 |
| **Marital Status** |  |  |  |  |  |
| Unmarried | 1.00 (ref) |  |  |  |  |
| Married | 1.292 | 0.523 | 0.527 | 0.584 | 2.857 |
| **Epilepsy Classification** |  |  |  |  |  |
| Generalized | 1.00 (ref) |  |  |  |  |
| Focal | 0.794 | 0.373 | 0.624 | 0.316 | 1.995 |
| **ASM prescribed prior to enrollment** |  |  |  |  |  |
| No | 1.00 (ref) |  |  |  |  |
| Yes | 3.091 | 1.363 | **0.011** | 1.302 | 7.337 |
| **No. ASMs prescribed** |  |  |  |  |  |
| Monotherapy | 1.00 (ref) |  |  |  |  |
| Polytherapy | 3.885 | 1.740 | **0.002** | 1.615 | 9.345 |

### Male comparator population

Among biologically male participants, there were 637 in the reproductive age range of 15 to 49 with a mean age of 29 (SD 9.7). Males were less likely to be married than their female counterparts (p<0.001) but had similar rates of developmental delay and distribution of epilepsy classification. For NCC, the distribution was also similar between females and male comparators in the same age range with 38% of males meeting probable or definite NCC criteria. For prescriptions, 506/637 (79.4%) of males were prescribed at least one ASM. Monotherapy was used in 457/506 (90.3%) and polytherapy in 49/506 (9.7%). Again, newer generation ASM prescriptions were exceedingly rare (2.3%). Like their female counterparts, most prescriptions were for “Possible Risk/Inadequate Data” category medications (88.1%) with the remainder of prescriptions in the “Established High Risk” category (11.9%). ASM risk profiles did not differ significantly between sex groups (p > 0.05) (Supplemental Table 1).

## Discussion

In this large, population-based epilepsy cohort from a *Taenia solium*–endemic region of Northern Peru, we observed that during the study period from 2006 to 2020, people of childbearing potential were predominantly treated with first-generation ASMs including carbamazepine, phenytoin, phenobarbital, and valproate, the ASMs reliably available in this region during this time. While phenytoin and carbamazepine are categorized as “Possible Risk/Inadequate Data,” valproate is “established high risk” for fetal malformations and adverse neurodevelopmental outcomes.^22–25^ Despite evidence supporting the use of newer generation agents like lamotrigine and levetiracetam for people with epilepsy who may become pregnant, the prescription patterns seen in our cohort reflect local formulary constraints. This is consistent with prior work in LMICs demonstrating that first-generation ASMs remain the backbone of epilepsy care, with newer, safer agents rarely accessible through public health systems.^12,26–28^ Our data extend these observations to a uniquely vulnerable group, people of childbearing potential living in an NCC-endemic region, where both seizure burden and reproductive risk are elevated.

Prenatal exposure to valproate is associated with a substantially increased risk of major congenital malformations, including neural tube defects, craniofacial anomalies, and cardiac malformations, with risk rising in a dose-dependent manner. In addition to structural defects, in utero valproate exposure is linked to long-term neurodevelopmental impairments, including reduced cognitive performance, language delays, and increased risk of autism spectrum disorder and attention-deficit/hyperactivity disorder also in a dose dependent manner. ^4,23,24,29^ In this cohort, while there is a clear preference for avoiding valproate where possible, still more than 12% of women of reproductive age are receiving valproate. The similarity in ASM prescription risk profiles between females of childbearing potential and males in the same age range suggests that reproductive status does not meaningfully influence prescribing decisions in this setting.

The social implications and financial cost of valproate use in pregnancy is staggering. The risk of major congenital malformations alone, the most overt risk, for all doses of valproate is over 9%. For doses of valproate 1450mg and over, the risk of major congenital malformation is over 25%.^30^ The cost and visibility of adverse neurodevelopmental outcomes in children of people taking valproate in pregnancy is less clearly seen, but no less impactful. From a financial perspective alone, it is estimated in the United States, that increased costs of special education and other medical needs for children with neurodevelopmental deficits attributable to in utero ASM exposure, total approximately $626 million dollars each year.^31^ This estimate does not include the costs of malformations and behavioral disorders (e.g., autism) nor the emotional and financial impact on the child’s family.

In our analysis, we identified ASM prescriptions prior to enrollment and polytherapy at intake as predictors of valproate use. The association with prior ASM prescription prior to enrollment and polytherapy are likely reflective of greater seizure severity or drug-resistance. Unfortunately, polytherapy, regardless of valproate inclusion in the ASM regimen, in of itself has been identified as an independent risk factor for adverse neurodevelopmental fetal outcomes.^32,33^

Despite the clear clinical importance of balancing seizure control with reproductive risk, there is limited guidance tailored to low-resource settings where newer generation ASMs like lamotrigine and levetiracetam are not readily available. Most guidelines are developed based on practice contexts where a range of newer ASMs can be accessed and monitored with reliable laboratory support.^4,5^ In contrast, in many LMICs, these newer agents are either unavailable or unaffordable for most.^26,28^

This lack of context-specific guidance for LMICs leaves clinicians and patients navigating highly complex decisions with limited supportive evidence to choose among the available medications that carry a greater teratogenic potential. There is currently no widely accepted, resource-stratified framework to support ASM selection in these circumstances, nor sufficient data on outcomes among people of childbearing potential treated with first-generation ASMs under real-world constraints.

While current guidelines from high income country major health policy leaders including the America Academy of Neurology (AAN), US Food and Drug Administration (FDA) and European Medicines Agency (EMA) make clear the importance of avoiding valproate in people of childbearing potential,^34–36^ even the WHO with its more global, and broadly inclusive mission still falls short of providing recommendations beyond this clear prohibition.^37^ The absence of guidance is likely due mostly to a scarcity of longitudinal, large-scale research conducted in LMICs that examines ASM effectiveness, safety, and reproductive outcomes where newer ASMs are not routinely used. As a result, clinicians frequently must rely on best clinical judgment, balancing seizure type, severity, and potential reproductive risk within the narrow range of medications available.

These findings highlight a need for pragmatic, context-aware guidance that supports ASM selection where choice is inherently limited. Such guidance would explicitly address scenarios in which newer generation ASMs are unavailable and offer risk mitigation strategies including guidance on reproductive health counseling. Strengthening clinical support tools, including simplified decision aids that incorporate medication availability, seizure type, teratogenic risk, and reproductive intentions, could help standardize care and improve shared decision-making.

Strengths of this study include its population-based design, large sample size, and systematic clinical characterization, including NCC status. However, several limitations should be acknowledged. We lack participant family planning data or understanding of pregnancy intentions. We also lack data on specific individualized provider decision-making parameters. There is also significant missingness among numerous co-variates, which particularly skews the representativeness of the sub-population used in the multivariate regression analysis, limiting interpretability (Table S3). Nonetheless, missingness is inherent to medical record data when leveraged as a secondary data source. Despite these limitations, these data are important to present given the scarcity of information capturing the experience of this epilepsy population.

Finally, this data is from 2006 to 2020 and there have been changes to ASM availability in Peru since this time. Notably, levetiracetam was evaluated for inclusion in Peru’s National Formulary in 2017 and incorporated within a complementary list for refractory epilepsy. In the most recent formulary from 2023, it remains a restricted therapy, requiring specialist justification and institutional approval.^38^ With this update, levetiracetam is now more widely available in Peru, but because of restrictions, its availability in rural settings remains limited, where access to specialty care and approval pathways is constrained. In rural Peru, treatment continues to include predominantly older, first-line ASMs.

In summary, this study reveals that in a *T. solium* endemic, resource-limited setting, ASM prescribing for people of childbearing potential is more likely shaped by medication availability and seizure control needs, rather than teratogenic risk profiles. Valproate and other older ASMs are widely used not because clinicians are unaware of reproductive risks, but because these agents are often the most effective and reliable treatment options available. This underscores the critical need for guidance that recognizes the realities of low-resource epilepsy care and supports clinicians and patients in making informed decisions that best balance seizure control with reproductive considerations.

## Data Availability

All data produced in the present study are available upon reasonable request to the authors

