## Supplemental Tables for "High-Risk Anti-Seizure Medication Use in Childbearing-Age People with Epilepsy in a *Taenia solium* Endemic Region"

Supplemental Materials

Table S1: Baseline characteristics of Male and Female Participants

| | Females No. (%) | Males No. (%) | P value |
|---|---|---|---|
| **Participants aged 15-49** | 685 | 637 | |
| **Average age (SD)** | 30.1 (9.9) | 29.1 (9.7) | 0.11 |
| **Married marital status** | 360 (52.6) | 226 (37.1) | **<0.001** |
| **History of developmental delay** | 72 (10.5) | 73 (11.5) | 0.48 |
| **Epilepsy Type** | | | |
| Generalized | 116 (16.9) | 115 (18.1) | 0.60 |
| Focal | 411 (60.0) | 359 (56.4) | |
| Unknown | 12 (1.8) | 12 (1.9) | |
| Missing | 146 (21.3) | 151 (23.7) | |
| **NCC Diagnosis** | 239 (34.9) | 243 (38.2) | 0.26 |
| **Duration of epilepsy** | | | |
| 0-10 years | 272 (39.7) | 273 (42.9) | 0.48 |
| >10 years | 310 (45.3) | 277 (43.5) | |
| Missing | 103 (15.0) | 87 (13.7) | |
| **ASM prior to recruitment** | 162 (23.6) | 158 (24.8) | 0.62 |
| **Number of ASMs prescribed at baseline** | | | |
| None | 138 (20.2) | 131 (20.6) | 0.78 |
| Monotherapy | 487 (71.1) | 457 (71.7) | |
| Polytherapy | 60 (8.8) | 49 (7.7) | |
| **First Generation ASM prescription*** | 538 (98.6) | 493 (97.4) | 0.30 |
| **ASM risk profile*** | | | |
| Low Risk | 1 (0.2) | 0 (0) | 0.56 |
| Possible Risk/Inadequate Data | 476 (87.0) | 446 (88.1) | |
| High Risk | 70 (12.8) | 60 (11.9) | |

* Denominator out of total participants prescribed ASMs at baseline (n=547 for females and n=506 for males)

**Table S2: Prescription counts by ASM before and after cohort recruitment for female participants of reproductive age**

| | ***Before*** | ***After*** |
|---|---|---|
| **Carbamazepine** | 82 | 365 |
| **Valproate** | 44 | 70 |
| **Phenytoin** | 33 | 113 |
| **Phenobarbital** | 4 | 29 |
| **Benzodiazepine** | 11 | 23 |
| **Lamotrigine** | 4 | 6 |
| **Levetiracetam** | 3 | 0 |
| **Topiramate** | 1 | 3 |
| **Other** | 1 | 1 |
| **Total # of prescriptions** | 326 | 613 |

Counts represent individuals prescribed medication(s) and will be greater than the total analytic population given the presence of polytherapy

**Table S3: Characteristics of Participants Included in the Regression Analysis (n=211) compared with those excluded due to missingness (n=336)**

| | Included n (%) | Excluded n (%) | P value |
|---|---|---|---|
| **Participants aged 15-49** | 211 | 336 | |
| **Average age (SD)** | 29.1 (9.7) | 29.8 (9.9) | 0.46 |
| **Married marital status** | 117 (54.1) | 159 (49.7) | 0.19 |
| **History of developmental delay** | 21 (13.2) | 37 (14.2) | 0.78 |
| **Epilepsy Type** | | | |
| Generalized | 39 (18.5) | 46 (20.3) | **0.01** |
| Focal | 172 (81.5) | 172 (75.8) | |
| Unknown | 0 (0) | 9 (4.0) | |
| **NCC Diagnosis** | 76 (39.0) | 120 (39.6) | 0.89 |
| **Duration of epilepsy** | | | |
| 0-10 years | 113 (57.7) | 124 (44.9) | **0.01** |
| >10 years | 83 (42.4) | 227 (55.1) | |
| **ASM prior to recruitment** | 112 (53.1) | 27 (8.0) | **<0.001** |
| **Number of ASMs prescribed at baseline** | | | |
| Monotherapy | 180 (85.3) | 307 (91.4) | **0.03** |
| Polytherapy | 31 (14.7) | 29 (8.6) | |
| **First Generation ASM Prescription** | 204 (96.7) | 334 (99.4) | **0.02** |
| **ASM risk profile** | | | |
| Low Risk | 1 (0.5) | 0 (0) | **<0.001** |
| Possible Risk/Inadequate Data | 169 (80.1) | 307 (91.4) | |
| High Risk | 41 (19.4) | 29 (8.6) | |
